# A comparison of knowledge-based dose prediction approaches to assessing head and neck radiotherapy plan quality

**DOI:** 10.1101/2024.01.24.24301485

**Authors:** Alexandra O. Leone, Mary Gronberg, Skylar S. Gay, Pavel A. Govyadinov, Beth M. Beadle, Tze Y. Lim, Thomas J. Whitaker, Karen Hoffman, Laurence E. Court, Wenhua Cao

**Affiliations:** Department of Radiation Physics, The University of Texas MD Anderson Cancer Center, Houston, Texas, USA; The University of Texas MD Anderson Cancer Center UTHealth Houston Graduate School of Biomedical Sciences, Houston, Texas, USA; Department of Radiation Oncology, Stanford University, Stanford, California, USA; Department of Radiation Oncology, The University of Texas MD Anderson Cancer Center, Houston, Texas, USA

## Abstract

**PURPOSE:** Recent studies demonstrate deep learning dose prediction algorithms may produce results like those of traditional knowledge-based planning tools. In this exploratory study, we compared 2D DVH-based knowledge-based planning tools and 3D deep learning-based approaches to assessing radiotherapy plan quality.

**METHODS:** Pre-validated 2D and 3D dose prediction models were applied to 58 patients with head and neck cancer treated under RTOG 0522 obtained from The Cancer Imaging Archive (TCIA). The 2D model was used to predict dose-volume histogram bands for seven organs at risk (OARs; brainstem, spinal cord, oral cavity, larynx, mandible, right parotid, and left parotid). A 3D dose prediction model was used to predict 3D dose distributions, based on computed tomography images, OAR contours, planning target volumes and prescriptions. The mean and D1% to the seven OARs for the 2D and 3D dose prediction models were compared. Further post predictive analysis was done to quantify the predicted 3D dose sparing for all normal tissues.

**RESULTS:** The two models predicted similar dose sparing to the OARs, with a mean difference of 1.4±5.5 Gy across all evaluated dose metrics. When looking at the sparing of non-OAR normal tissue regions, the 3D model predicted a mean dose reduction to normal tissue regions of 6.4±3.0 Gy when compared with the clinical dose.

**CONCLUSION:** 2D and 3D dose predictions are comparable at predicting dose reductions to OARs. The 3D approach allows for dose visualization, which may support further sparing of normal tissues not typically drawn as OARs on head and neck plans.

## 1 INTRODUCTION

Peer review of radiotherapy plans aims to identify suboptimal plan quality prior to delivery of therapy. However, suboptimal radiotherapy plans continue to make it through quality assurance undetected, with one study demonstrating that 45% of errors made it past the plan review process.[1] Furthermore, the same study showed that the effectiveness of the peer review process degrades as a peer-review meeting progresses.[1] Decision support tools could improve the efficiency and effectiveness of this peer-review process.

This notion has emphasized the importance of continued clinical efforts to improve the consistency and quality of radiotherapy planning using a variety of planning tools. [2-9] These include traditional knowledge-based planning tools that predict dose-volume histograms (DVHs), and deep learning-based tools, that predict dose distributions on computed tomography images for later DVH calculation. The more traditional knowledge-based planning tools typically use a library of plans from previously treated patients and develop models associating geometric features with their corresponding dosimetry to predict possibly achievable dosimetry for a new patient. This type of knowledge-based planning is commercially used in the RapidPlan tool (Varian Medical Systems, Palo Alto, CA) where a predicted dose range is estimated on the DVH for specific organs and/or targets at the time of plan optimization.

Knowledge-based planning tools such as this, are beneficial in demonstrating what doses can reasonably be achieved for each OAR in a specific patient plan. On the other hand, deep learning focused works can predict entire dose distributions as opposed to organ and/or target specific values. This method has proven beneficial in not only successfully predicting the dose for a variety of disease sites (Head and Neck [3,4], Lung [5], Prostate [6]), but also demonstrating what potential tradeoffs may result should an OAR be flagged for further sparing. Although both types of KBP were primarily developed to provide optimization objectives, these tools can also be used for assessing patient-specific plan quality.

Previous work by Ahn et al aimed to determine the difference in performance between a deep learning prediction model and conventional knowledge-based planning using left-breast clinical cases. Both models were generated using the same dataset and evaluation was done using the absolute dose difference error, DVH, 2D gamma index and iso-dose dice similarity coefficient. Their work found the doses predicted by deep learning were superior to the knowledge-based planning predictions for their data. [7] However, this study was limited as it was internally evaluated with 10 patients and 4 structures per patient. Further work is needed to look at different treatment sites and more structures.

In this exploratory analysis, we compared the ability of a DVH-based (2D) knowledge-based planning model [2] and a deep learning-based 3D dose prediction model[3,4] to predict optimal dose distributions for 58 patients and 7 organs at risk with head and neck cancer in a phase 3 trial (Radiation Therapy Oncology Group [RTOG] 0522) using data available on The Cancer Imaging Archive.[10-12] To the best of our knowledge, this is the first study to directly compare these two methods with a single independent head and neck data set. We demonstrated the potential use of these plan optimization tools for quality assessment with application to automating quality assurance for clinical trial plans and support of peer review of routine clinical plans.

## 2 MATERIALS AND METHODS

The widely tested traditional KBP system uses organ at risk (OAR) and planning target volume geometry to predict a range of DVH estimates for a given OAR.[2] Deep learning–based 3D dose prediction is a more recently developed knowledge-based planning (KBP) approach that predicts an entire dose distribution. These models typically receive the same planning inputs as manual planners, including patient computed tomography (CT) images, normal tissue and target contours, and prescriptions. [3,4]

### 2.1 Patient data

A DICOM data set for 111 head and neck cancer patients in the RTOG 0522 trial was obtained from The Cancer Imaging Archive. The RTOG 0522 trial was a randomized phase 3 trial of concurrent accelerated radiotherapy and treatment with cisplatin compared with concurrent accelerated radiotherapy and treatment with cisplatin and cetuximab for stage III and IV head and neck carcinoma. The patients received radiotherapy for cancers in the hypopharynx, oropharynx, or larynx region from 2005 to 2009.[10-12] Of the 111 patients, 27 were excluded because they underwent 3D conformal radiotherapy, and 26 were excluded because of missing CT slices or dose grids much larger than those in the data the DVH-based and/or deep learning-based model were trained with.

For each of the remaining 58 intensity modulated radiation therapy (IMRT) plans, seven OARs were assessed when available—brainstem, spinal cord, larynx, mandible, right parotid, left parotid, and oral cavity— using the mean dose (larynx, parotids, and oral cavity) and D1% dose (brainstem, spinal cord, mandible, and oral cavity). D1% was calculated as a surrogate to maximum dose to account for differences in treatment planning system calculation of max dose.

### 2.2 Model Information

The DVH predictions were generated using a previously validated 2D head and neck cancer model trained using 25 volumetric modulated arc therapy plans created by a fully automated TPS, the Radiation Planning Assistant. This system can generate consistent, high-quality volumetric modulated arc therapy plans for head and neck cancer patients as described previously.[13] We previously demonstrated that this model could predict insufficient dose sparing in head and neck plans also identified by radiation oncologists.[2] Deep learning-based 3D dose predictions were created using a validated internal head and neck cancer model. The model was trained, validated, and tested with head and neck volumetric modulated arc therapy plans created with the 2D model. [3,4] More information on these models, the datasets used to train them, and their validation for plan quality assurance can be found in reference 2, 3 and 4.

### 2.3 Evaluation of the DVH-based KBP prediction model

For each patient, dose predictions were created using the 2D DVH-based KBP prediction model for each OAR available in the clinical plan. The average predicted dose value was determined for each OAR based on the upper and lower dose estimate for a given clinical objective (e.g., mean parotid dose) and compared with the clinical value. This process was completed for all 58 patients in the cohort.

### 2.4 Evaluation of deep learning–based dose prediction

The dose distribution for each patient was predicted and visualized using a RayStation TPS (RaySearch Laboratories, Stockholm, Sweden). The same dose metrics - mean dose (larynx, parotids, and oral cavity) and D1% dose (brainstem, spinal cord, mandible, and oral cavity) - were then evaluated and compared with the DVH predictions and clinical values.

A limited body contour was created to quantify dose falloff from the planning target volume to both OARs and non-OAR normal tissues. This was created using the RayStation system by subtracting the expanded target region (all planning target volumes with uniform expansion of 2 cm) from the body contour, and then limiting the resulting structure to the same superior and inferior bounds as the expanded target region. The mean dose delivered to this structure was evaluated and used as an indicator of suboptimal dose falloff from the targets.

We conducted a paired two-sided Student’s *t*-test [14] to assess the equivalence of 2D and 3D dose prediction values for each of the seven OARs with a p-value significance threshold of > 0.05 indicating no statistically significant difference between the two methods.

## 3 RESULTS

The comparison of the two dose prediction models resulted in the lowest t value being 0.083 (*p* = 0.93) and the highest t value being 6.89 (*p* < 0.001). The mean (± SD) overall difference in OAR dose sparing between the 2D and 3D models was 1.4 ± 5.5 Gy (Figure 1).

**FIGURE 1.**
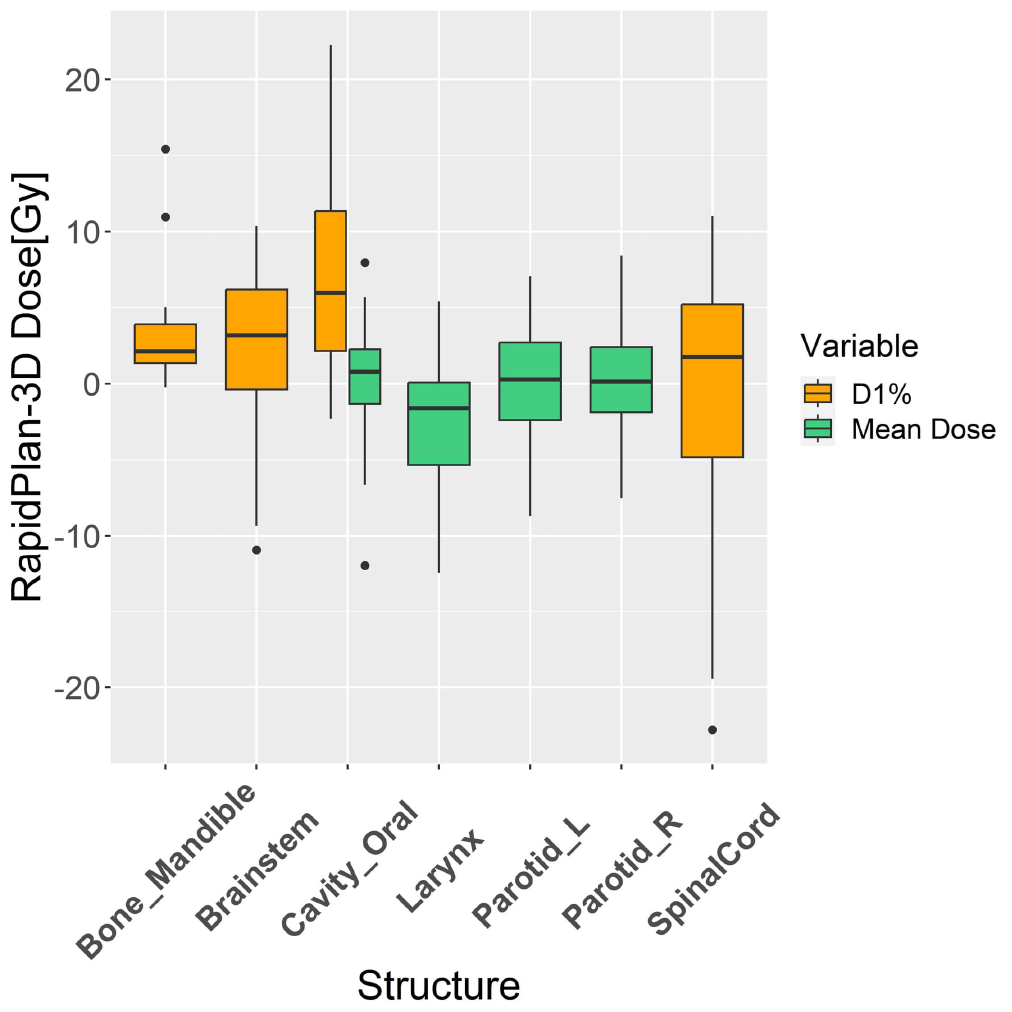
Boxplot of the differences in predicted dose between the 2D KBP versus the 3D deep-learning based dse prediction model.

We calculated the absolute difference between the predicted and clinical values for each patient’s OAR dose metric using the DVH-based 2D and deep learning–based 3D dose prediction models (Figure 2). As demonstrated by the median values in the box plots in Figure 2, for most of the structures in the study patients, the 2D and 3D dose predictions resulted in similar dose reduction values. For example, the mean left parotid dose was predicted to be spared by an additional 5.6 ± 7.0 Gy when using the 2D dose prediction model and 5.6 ± 7.2 Gy when using 3D dose prediction model. Likewise, the spinal cord D1% was predicted to be spared by an additional 5.8 ± 6.5 Gy with the 2D dose prediction model and 6.6 ± 8.1 Gy with the 3D dose prediction model. Reviewing the individual dosimetric values, the agreement between the 2D predicted and 3D deep learning based predicted dose falls within 5 Gy for most patients (Figure 3). This was expected, as we trained the 2D and 3D dose prediction models with similar quality data.

**FIGURE 2.**
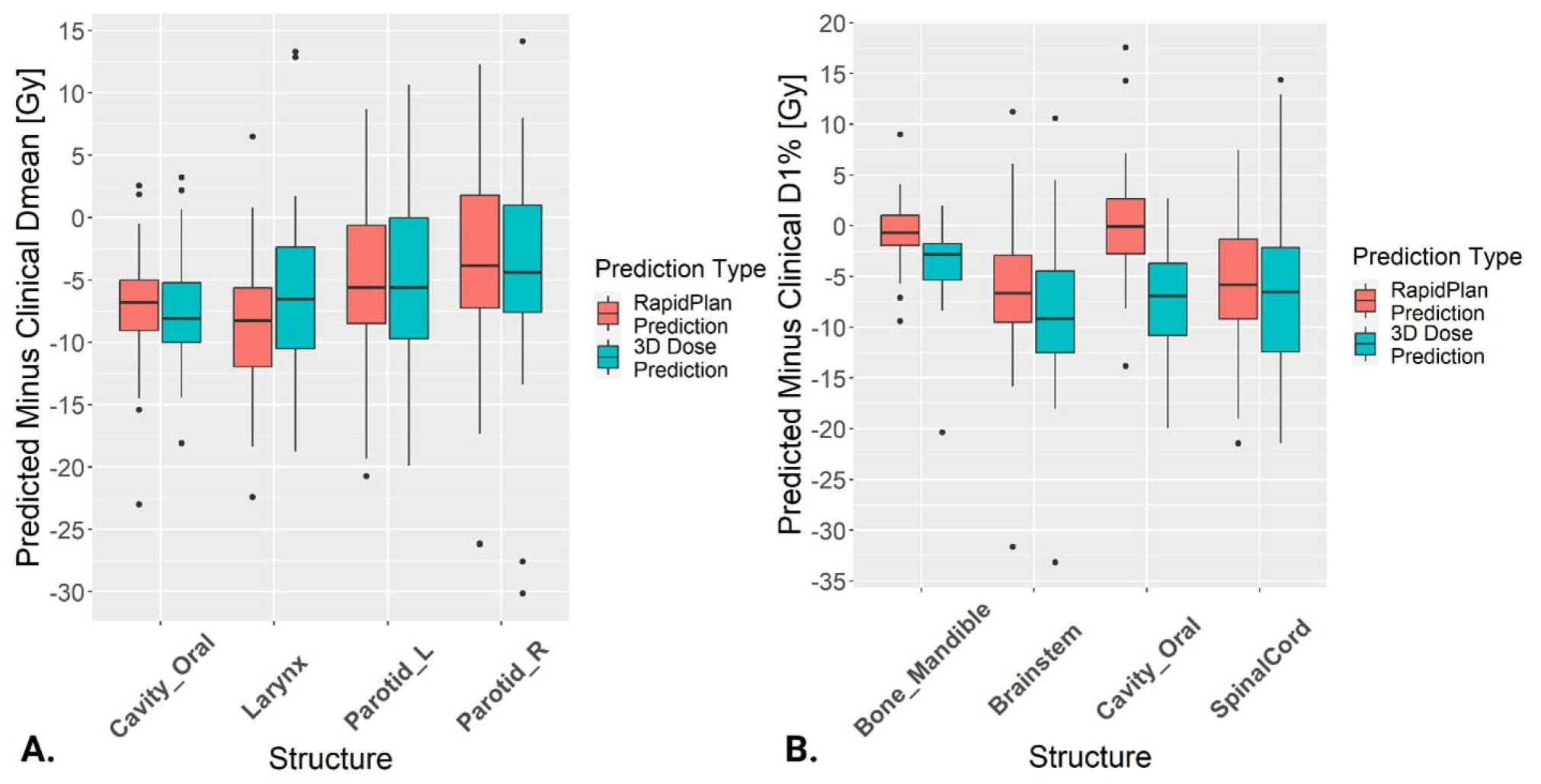
Box plots of the differences between the predicted (2D and 3D Dose Prediction) and clinical dose reduction values for (A) mean dose OARs and (B) D1% OARs.

**FIGURE 3.**
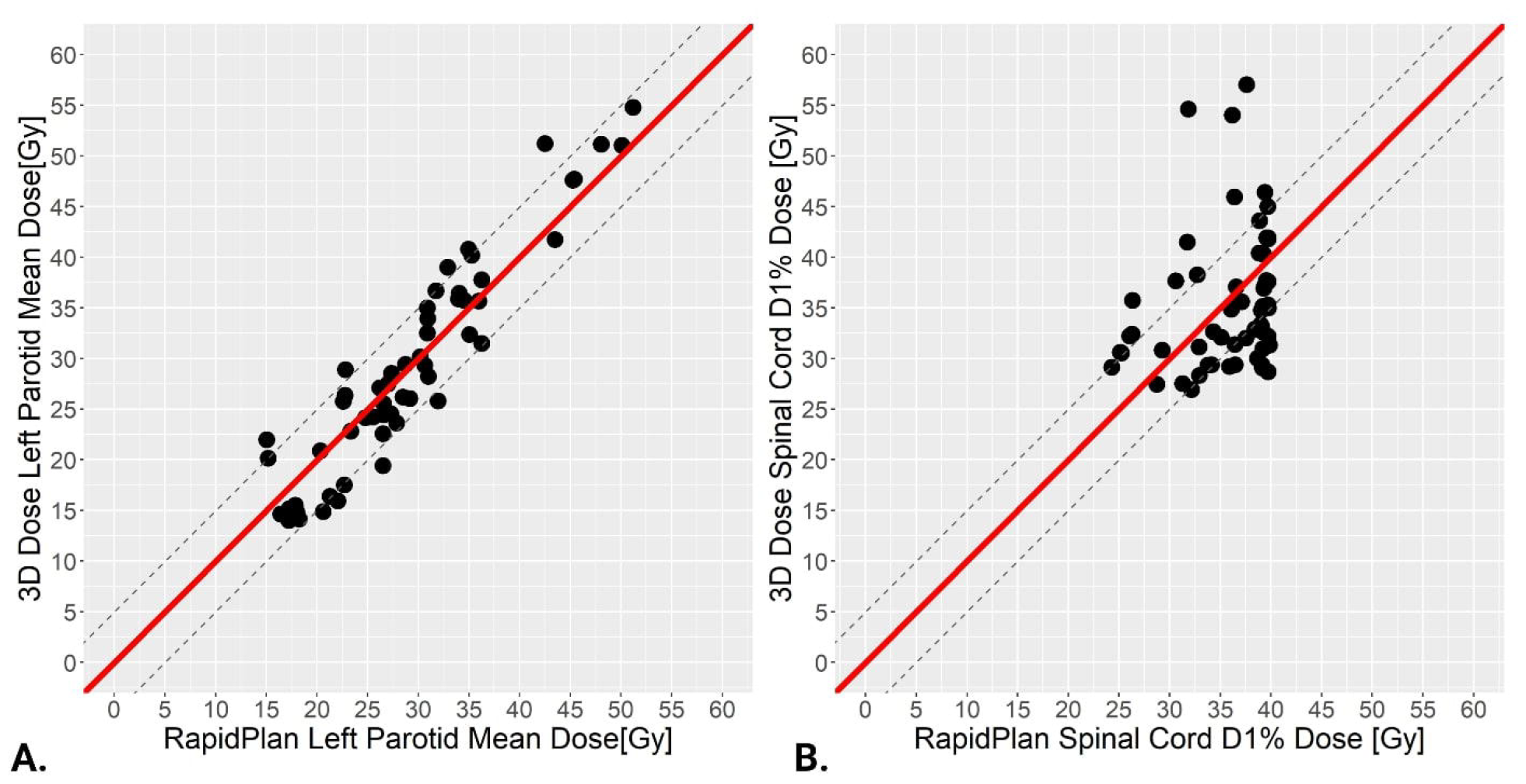
Dot plot comparison of the 2D and 3D dose prediction DVH values per patient. The dashed lines in both plots represent 5 Gy intervals for (A) Left parotid mean dose and (B) Spinal cord D1%.

An example of a patient with large differences in the predicted DVH and 3D dose distribution is shown by sample patient 1 in Figure 4. This patient had a large target volume near the spinal cord. The 2D model predicted an average D1% of 37.6 Gy for this patient. In contrast, the 3D dose prediction model predicted a D1% of 57 Gy, which exceeds the clinical tolerance of 45 Gy per standard planning guidelines. This case is not representative of the data that the 3D dose prediction model was trained with, as the target volume is much larger and closer to the spinal cord than are the target volumes in the cases in the training data.

**FIGURE 4:**
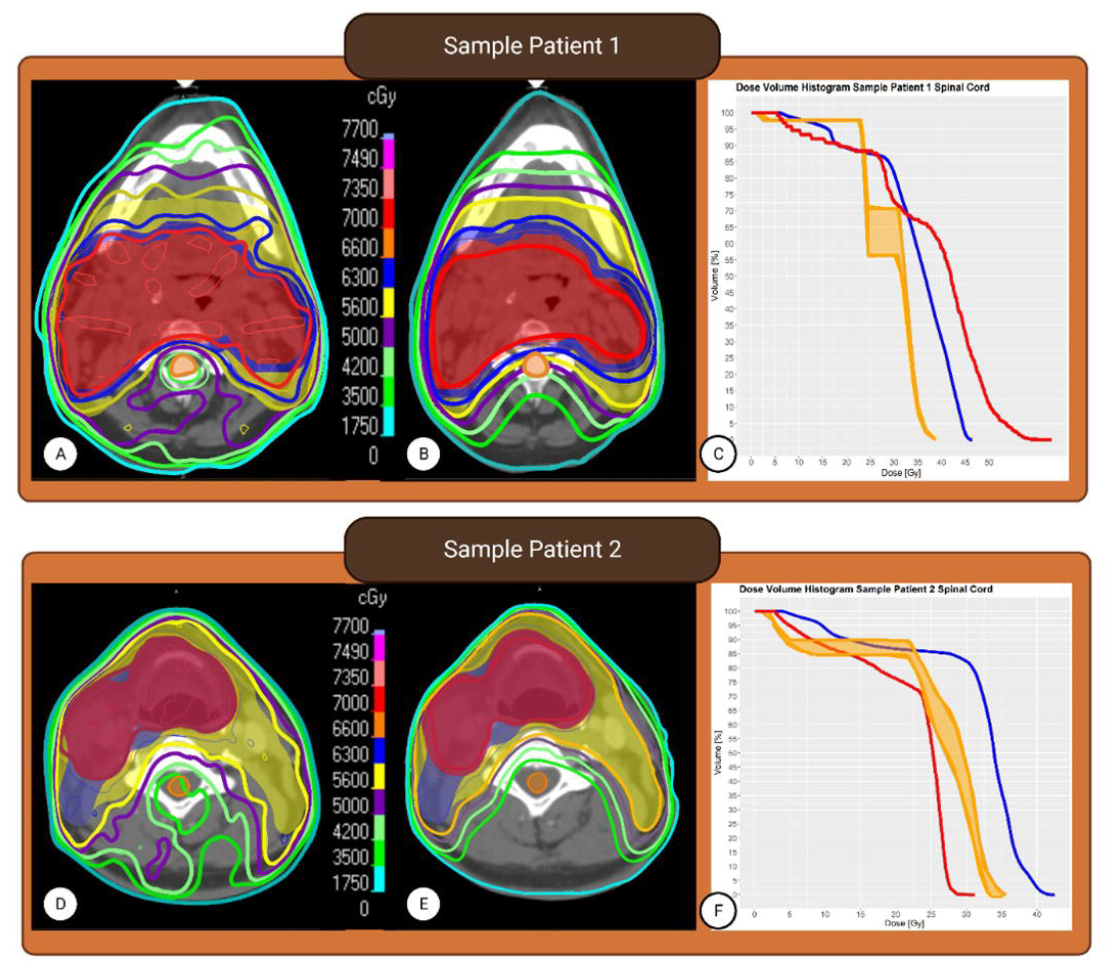
Two sample head and neck cancer patients in our cohort. (A and D) Dose distributions and spinal cord contour for the clinical plans. (B and E) Dose distributions as predicted by the deep-learning model with the clinical spinal cord contour. (C and F) The spinal cord DVHs for 3D dose predictions (red), 2D dose predictions (orange), and clinical plan (blue).

We evaluated the absolute difference in mean dose to the limited body structure, observing a predicted mean dose reduction of 6.4 ± 3.0 Gy when using 3D dose prediction as compared with the clinical plans. As seen with our sample patient 2 in Figure 4, both the 3D distributions and DVHs demonstrated potential dose sparing in the spinal cord region as well as the posterior region of the brain.

## 4 DISCUSSION

The present work suggests that both 2D and 3D dose prediction techniques can predict dose sparing and identify suboptimal dose distributions. Both techniques provide similar predictions of dose metrics, as seen in similar comparison studies in the literature.[15] Considering this, the quality assurance measure of flagging cases with suboptimal doses could be carried out with either technique. Demonstrating the suitability of both models for plan quality assessment was important to providing flexibility of choice to users who may prefer one dose prediction technique over the other. In addition, the normal tissue structure provides a quantitative measure to demonstrate that a 3D dose prediction model provides a unique lens of plan optimization apart from flagging subpar cases based on structure clinical objective performance by enabling the user to visualize a dose not easily interpreted on a DVH (e.g., falloff regions from PTVs, locations of hot and cold spots). The ability to visualize a dose in this manner can be useful in determining the dosimetric trade-off to surrounding normal tissues by OAR dose sparing, if any, providing an added layer of validity to these predictive measures. This additional support can persuade clinicians to make improvements to current treatment plans that they would otherwise be skeptical about.

We observed that the 2D model was more robust than the 3D model in predicting doses in cases that were not representative of the data used to train the models. For example, in cases in which the spinal cord was near the target, the 3D model predicted a D1% that exceeded tolerances. Prior to implementing 3D dose prediction for plan quality assurance, the predicted doses should also be checked with a scorecard to ensure that the dose to normal tissues are meeting clinical tolerances.

While more robust, 2D predictions can also be limited to the OARs present at planning. This means any prediction made by the 2D model is based solely on the structures that were planned and does not consider any OARs not available in the model. For example, swallowing structures are not often contoured but have been identified as regions with strong dose-effect relationships correlating with dysphagia and reduced quality of life [16,17]. Utilizing a DVH-based model, predictions are not able to be generated and a new model must be created for these structures. However, as deep learning-based predictions are mainly target volume dependent, a prediction can be generated regardless of present structures and would only require additional training if there is a desire to reduce dose to these new structures more than is predicted. This is emphasized in the present study with the use of the limited body contour. While we are unable to predict dose to our limited body structure utilizing the 2D model, we can see using the 3D model that dose to the posterior region behind the spinal cord and swallowing structure region lateral to the esophagus is able to be reduced without specifically identifying these areas with contours (Figure 5).

**FIGURE 5:**
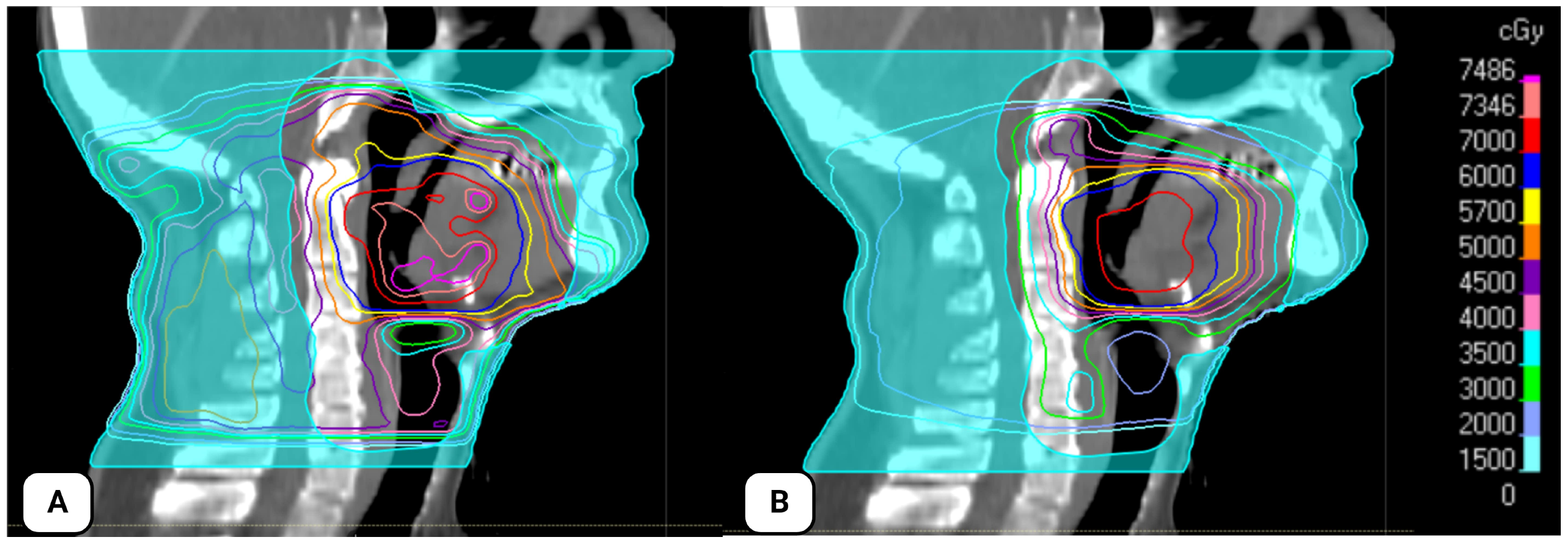
Sample patient in our cohort with visualized limited body contour and dose distributions. (A) Clinical dose distribution within limited body contour (blue). (B) Dose distributions as predicted by the deep-learning model within the limited body contour (blue).

Based on our findings, both 2D and 3D dose prediction methods are comparable in identifying potential areas of plan quality improvement. Plans that are atypical or where there are concerns regarding specific OARs, the DVH-based 2D prediction could be used. Plans that are standard with no clear areas of concern or with uncommon structures, may benefit from a predicted dose distribution. Visualizing the predicted dose distribution enables a planner to see what the potential dosimetric trade-off may be, if any, for achieving additional sparing of an OAR. This can provide an added layer of support for these predictive measures and thus may be more useful than DVH prediction in persuading clinicians to change a suboptimal treatment plan.

A limitation of this study is the technology used to create and deliver the studied radiotherapy plans. As these patients received treatment in the early days of intensity-modulated radiotherapy (2005-2009), and there has been considerable improvement in the planning and delivery of IMRT since that time. Since the plans were delivered over a decade ago with available technology and approaches, we cannot necessarily claim these plans should have better quality than what was clinically delivered.[18-20] However, this does not affect our ability to evaluation of the ability dose prediction models, such as DVH-based 2D and deep learning-based 3D dose predictions, in their ability to flag subpar treatment plans. Future studies of this work may include new models designed to increase the robustness of the 3D dose prediction model to accommodate the use of unfamiliar plans, evaluation in other disease sites, and studies using a larger cohort with more recent radiotherapy plans.

In conclusion, knowledge-based planning tools such as 2D DVH prediction and deep learning–based 3D dose prediction show promise to be used for automatic radiotherapy plan quality evaluation. The current implementation of a 2D DVH-based prediction was more robust in handling unfamiliar plans, while the 3D dose prediction model allowed for visualization of the predicted dose distribution to facilitate both OAR and non-OAR normal tissue sparing. Ultimately, plan quality could be evaluated with either tool dependent on clinical availability of these tools.

## Data Availability

All treatment plan data is available online at

https://wiki.cancerimagingarchive.net/display/Public/Head-Neck+Cetuximab#688455175bb463d4a54419a93c4112e24902bbc

